# Implementing a context-augmented large language model to guide precision cancer medicine

**DOI:** 10.1101/2025.05.09.25327312

**Authors:** Hyeji Jun, Yutaro Tanaka, Shreya Johri, Filipe LF Carvalho, Alexander C. Jordan, Chris Labaki, Matthew Nagy, Tess A. O’Meara, Theodora Pappa, Erica Maria Pimenta, Eddy Saad, David D Yang, Riaz Gillani, Alok K. Tewari, Brendan Reardon, Eliezer Van Allen

## Abstract

The rapid expansion of molecularly informed therapies in oncology, coupled with evolving regulatory FDA approvals, poses a challenge for oncologists seeking to integrate precision cancer medicine into patient care. Large Language Models (LLMs) have demonstrated potential for clinical applications, but their reliance on general knowledge limits their ability to provide up-to-date and niche treatment recommendations.

To address this challenge, we developed a RAG-LLM workflow augmented with Molecular Oncology Almanac (MOAlmanac), a curated precision oncology knowledge resource, and evaluated this approach relative to alternative frameworks (i.e. LLM-only) in making biomarker-driven treatment recommendations using both unstructured and structured data. We evaluated performance across 234 therapy-biomarker relationships. Finally, we assessed real-world applicability of the workflow by testing it on actual queries from practicing oncologists.

While LLM-only achieved 62–75% accuracy in biomarker-driven treatment recommendations, RAG-LLM achieved 79–91% accuracy with an unstructured database and 94–95% accuracy with a structured database. In addition to accuracy, structured context augmentation significantly increased precision (49% to 80%) and F1-score (57% to 84%) compared to unstructured data augmentation. In queries provided by practicing oncologists, RAG-LLM achieved 81–90% accuracy.

These findings demonstrate that the RAG-LLM framework effectively delivers precise and reliable FDA-approved precision oncology therapy recommendations grounded in individualized clinical data, and highlight the importance of integrating a well-curated, structured knowledge base in this process. While our RAG-LLM approach significantly improved accuracy compared to standard LLMs, further efforts will enhance the generation of reliable responses for ambiguous or unsupported clinical scenarios.

## Introduction

Identifying therapeutically targetable molecular alterations to guide treatment options is a key component of precision cancer medicine. However, the growing complexity and volume of regulatory approvals for such therapies make it increasingly challenging for clinicians to stay up to date with relevant clinicogenomic relationships^1^. Tracking these approvals often requires navigating multiple scattered sources, including electronic health records (EHRs), NCCN guidelines, PubMed, and internal emails. Moreover, certain approvals may occur with limited or no publicity, further delaying awareness. This knowledge gap may hinder the timely implementation of new approvals into clinical practice, especially for clinicians not well-versed in cancer genomics^2^.

In recent years, the advancements in Large Language Models (LLMs) for addressing similar challenges across clinical medicine have gained attention. Studies have demonstrated the capability of LLMs in tasks such as patient-to-clinical trial matching^3–5^, performing clinical summarization tasks^6^, and achieving physician-level performance on medical board examinations^7^. There is also emerging interest in their potential application to support clinical decision-making in precision oncology^8–10^.

Despite these advancements, LLMs have limitations when handling niche and constantly evolving queries, particularly in the field of oncology^11^. These challenges are due to insufficient domain-specific training and reliance on potentially outdated data, which raises concerns about the accuracy and relevance of their output^12^. This issue is especially critical given these strategies guide oncologists in clinical decision-making. Furthermore, the overwhelming variety of ever-growing available LLMs complicates their adoption, as benchmarking them for specific use cases can be both time-consuming and computationally demanding.

Efforts to address these limitations are an active area of research^13^. Notably, Retrieval-Augmented Generation (RAG) has emerged as a promising approach, dynamically retrieving relevant information from external domain-specific databases to supplement an LLM’s general knowledge without modifying its internal weights^14–17^. In precision oncology, expertly curated databases such as Molecular Oncology Almanac (MOAlmanac), OncoKB, CIVIC, and MyCancerGenome, are used to guide oncologists treatment options that may be available for diverse genomic and disease indications^18–21^. As such, we hypothesized that LLMs augmented with input from expertly curated databases could specifically enhance clinical decision support, particularly for practicing medical professionals in underserved settings^22,23^.

In this study, we introduce a RAG-LLM approach that enables accurate and approval-derived therapy recommendations based on patients’ genomic biomarkers, disease type, treatment history, and other clinically relevant information for treatment planning (Figure 1). Our RAG-LLM method leverages the MOAlmanac, an expert-curated clinicogenomic interpretation database that compiles the latest precision oncology knowledge regarding relationships between molecular features and clinical actionability^18^, to improve queries of retrieving appropriate FDA-approved biomarker-based oncology therapies. We evaluate this approach on a synthetic dataset, finding that it retrieves the approved therapeutic option based on the provided clinical information with high accuracy. We additionally benchmark this approach on real-world questions provided by practicing oncologists, finding that this approach accurately identifies approved therapies for different diagnoses, clinical histories, and known genomic alterations.

**Figure 1.**
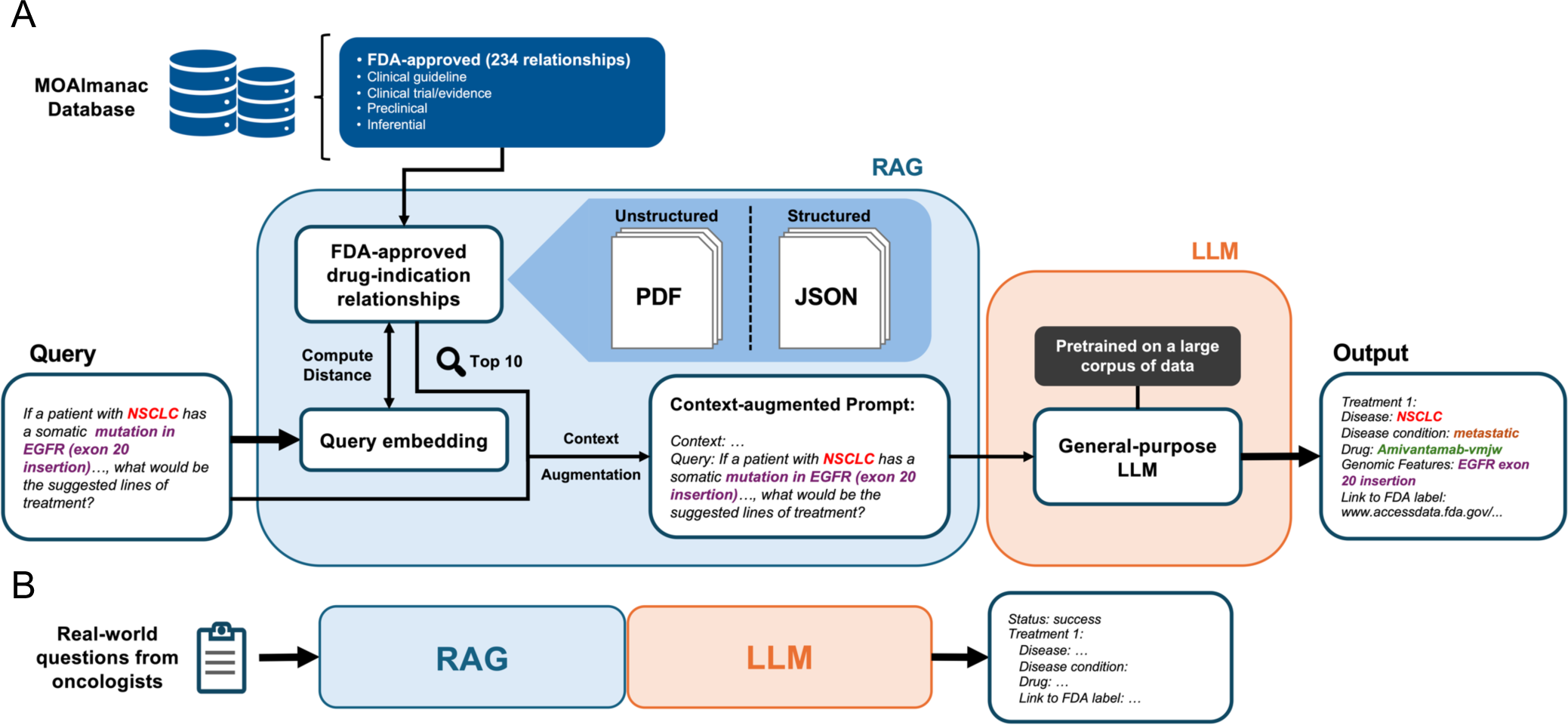
Schematic representation of the RAG-LLM precision oncology workflow for biomarker-driven treatment recommendations. **A.** Overview of the RAG-LLM precision oncology workflow. The RAG framework retrieves the top 10 most similar matches from the context database using Euclidean distance between query and context embeddings. To evaluate the impact of dataset structure on biomarker-driven treatment recommendations, we compared the performance of RAG-LLM with unstructured data augmentation versus that of RAG-LLM using structured data. The resulting context-augmented query was then fed into a general-purpose LLM for inference. **B.** To test real-world applicability of the RAG-LLM workflow, actual clinical queries collected from practicing oncologists were input into the RAG-LLM workflow.

## Results

### Prompt optimization and benchmarking of LLMs

Given the critical role of prompt optimization in enhancing LLM performance, we first evaluated whether designing a specific prompt structure would optimize the accuracy of a representative LLM (Mistral Nemo 12B) in accurately retrieving FDA-approved biomarker-based (“precision”) oncology therapies^24^. We tested four distinct prompt strategies: (1) a single instruction (referred to as the “basic prompt”), (2) a basic prompt with a condition statement restricting results to FDA-approved drugs only, (3) a basic prompt with a defined system role, and (4) a combination of the second and third additional statements (Table 1). A desired output format was specified in all strategies using a JSON-style schema. All evaluated queries were formulated using the structured data from MOAlmanac (Table S1–2). To compare prompt effectiveness, we measured partial match accuracy, defined as the proportion of retrieved therapies that matched ground-truth FDA-approved therapies (Methods). Among the four strategies, the basic prompt demonstrated the highest accuracy in retrieving FDA-approved therapies (Figure 2A). The basic prompt achieved an accuracy of 69.2%, outperforming the second (53.4%), third (64.5%), and fourth (51.3%) strategies.

**Figure 2.**
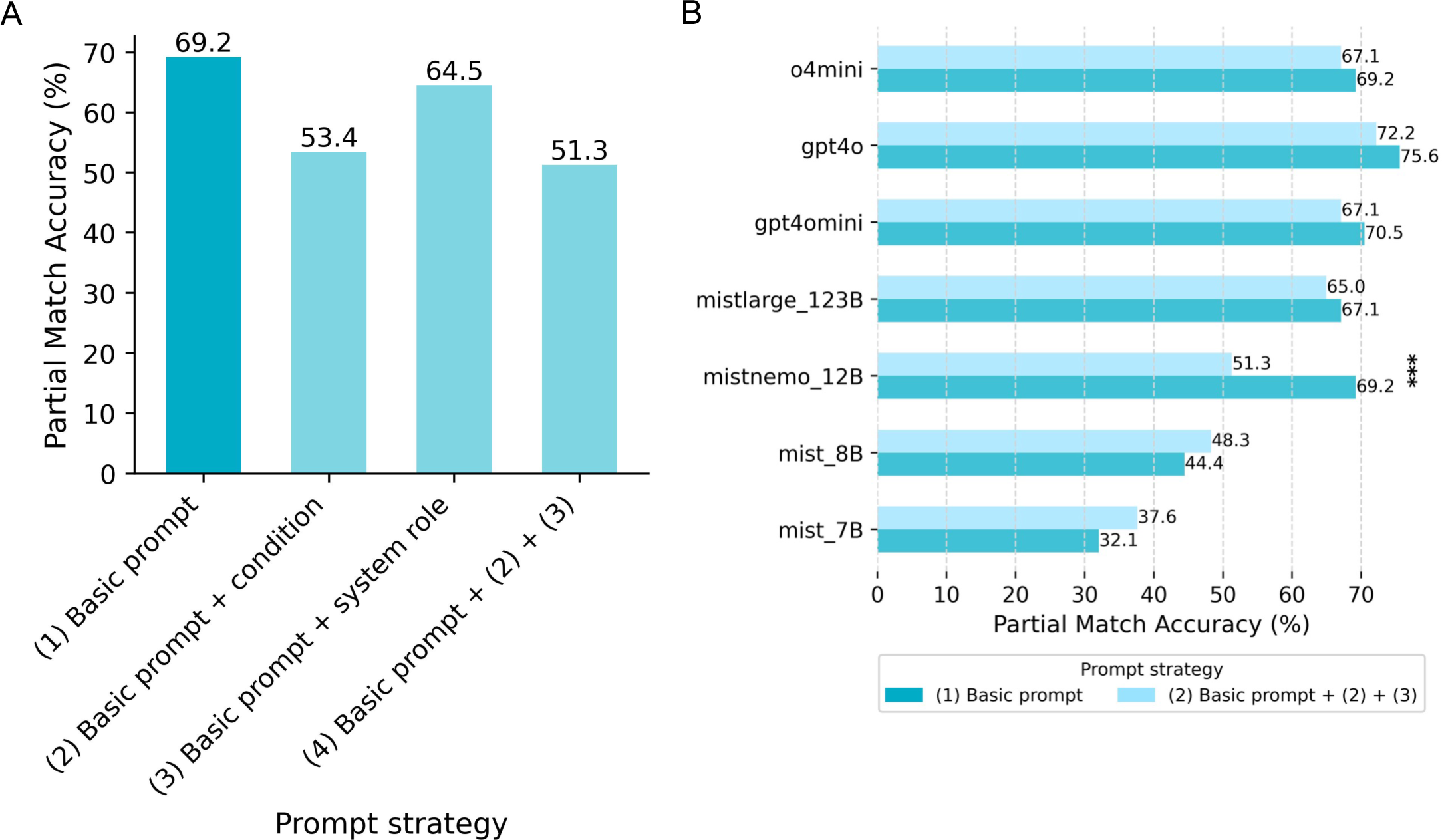
Performance of LLMs across various prompt engineering strategies. **A.** Partial match accuracies of Mistral NeMo 12B using different prompt strategies. **B.** Partial match accuracies of LLMs with varying sizes across basic and combined prompt strategies. Accuracies between the basic and combined prompt strategies were compared using McNemar’s test. P-value significance is shown if the p-value is less than 0.05 (p < 0.001: ***, p < 0.01: **, p < 0.05: *); otherwise, it is not displayed.

**Table 1.** Strategies tested for prompt design optimization.

| Strategy | Prompt |
| --- | --- |
| (1)<br>Basic prompt | Please provide each treatment as a json format with the following JSON schema.<br>`{{ "Treatment 1": {{ "Disease Name": , "Disease Phase or Condition": , "Drug Name": , "Prior Treatment or Resistance Status": , "Genomic Features": "Link to FDA-approved Label": }} }}` Query: {prompt} |
| (2)<br>Scope-limiting prompt | Please only provide the therapies that are FDA-approved for the provided genomic biomarkers. Please provide each treatment as a json format with the following JSON schema.<br>`{{ "Treatment 1": {{ "Disease Name": , "Disease Phase or Condition": , "Drug Name": , "Prior Treatment or Resistance Status": , "Genomic Features": "Link to FDA-approved Label": }} }}` Query: {prompt} |
| (3)<br>System role prompt | You are a helpful chat bot specialized in suggesting FDA-approved drugs to treat cancer. Please provide each treatment as a json format with the following JSON schema.<br>`{{ "Treatment 1": {{ "Disease Name": , "Disease Phase or Condition": , "Drug Name": , "Prior Treatment or Resistance Status": , "Genomic Features": "Link to FDA-approved Label": }} }}` Query: {prompt} |
| (4)<br>Combination prompt | You are a helpful chat bot specialized in suggesting FDA-approved drugs to treat cancer. Please only provide the therapies that are FDA-approved for the provided genomic biomarkers. Please provide each treatment as a json format with the following JSON schema.<br>`{{` |
|  | "Treatment 1": {{<br>"Disease Name": ,<br>"Disease Phase or Condition": ,<br>"Drug Name": ,<br>"Prior Treatment or Resistance Status": ,<br>"Genomic Features":<br>"Link to FDA-approved Label":<br>}}<br>}}<br>Query: {prompt} |

The superior performance of the basic prompting strategy was also consistently observed in other LLMs, particularly those of larger sizes (n = 7 LLM models tested; Figure 2B). Among all LLM models tested, GPT-4o achieved the highest accuracy. Based on these results, we selected the basic prompt and used GPT-4o for all subsequent evaluations in the RAG-LLM workflow, where relevant information from an external database was integrated into the prompt alongside the user-provided query.

### RAG-integrated LLM outperforms LLM in therapy predictions

We next hypothesized that structured data augmentation would improve the accuracy and reliability of LLM for FDA-approved therapy predictions compared to unstructured data. We thus evaluated GPT-4o’s performance using both unstructured and structured data formats. The unstructured dataset was constructed by extracting the ‘Indication and Usage’ section from drug label text, with a median length of 162 tokens (Interquartile range [IQR]: 127–240 tokens; Figure S1A, Table S3). In contrast, the structured dataset consisted of manually curated therapy-biomarker relationship pairs, with a median length of 181 tokens (IQR: 165–204 tokens; Figure S1B, Table S4). Both approaches were tested on 234 synthetic prompts derived from single entities in MOAlmanac, and accuracy was evaluated against ground-truth therapies (Methods).

Without RAG-provided context, model accuracy ranged from 62–75%. However, augmenting the model with unstructured text data significantly improved performance, increasing accuracy to 79–91% (exact match: χ^2^(1) = 153.38, p = 1.27 x 10^-34^; partial match: χ^2^(1) = 129.20, p = 1.23 x 10^-29^; McNemar’s test on results pooled from five repeated runs) (Figure 3A). Integrating structured data further enhanced performance compared to unstructured data augmentation, yielding an accuracy of 94–95% (exact match: χ^2^(1) = 103.80, p = 2.98 x 10^-24^; partial match: χ^2^(1) = 16.59, p = 4.64 x 10^-5^; McNemar’s test on results pooled from five iterations). In addition to accuracy, structured data augmentation markedly improved other key performance metrics (Figure 3B). Specifically, precision, recall, F1-score, and specificity all increased, with precision and F1-score significantly improving by approximately 50–60% relative to the unstructured data augmented model (mean precision: 49% to 80%, W = 15.0, p = 3.12 x 10^-2^; mean F1-score: 57% to 84%, W = 15.0, p = 3.12 x 10^-2^; one-sided Wilcoxon signed-rank test comparing model performance across five iterations). These results demonstrate the role of the RAG approach in enhancing the model’s ability to provide more precise and reliable FDA-approved therapy predictions, with structured data augmentation further optimizing performance.

**Figure 3.**
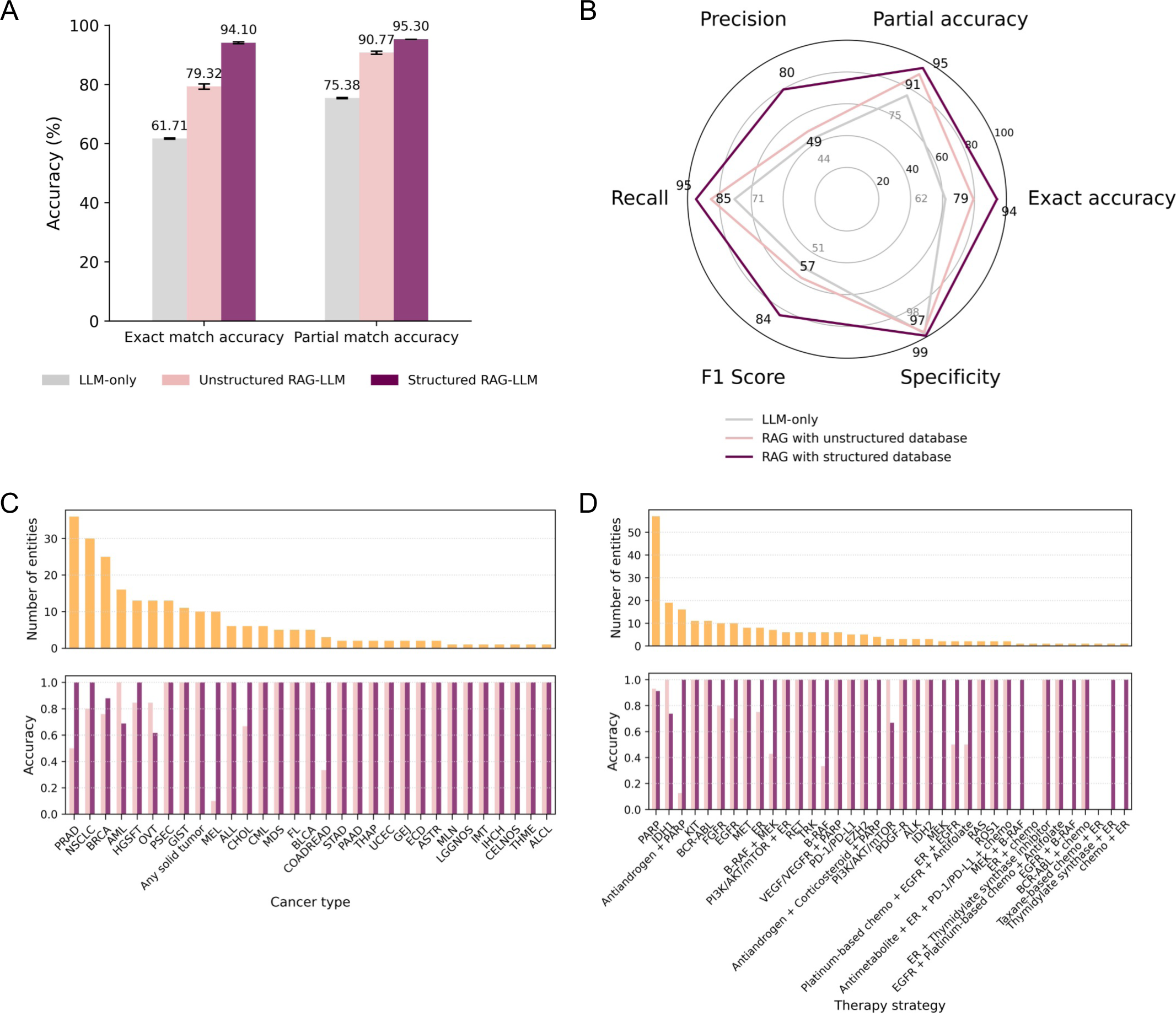
Enhancement through RAG using unstructured and structured datasets. **A.** Exact match and partial match accuracies from LLM-only and RAG-LLM augmented with unstructured and structured datasets. **B.** Average precision, recall, F1-score, specificity and accuracies from RAG-LLM with unstructured and structured data augmentation. **C.** Top: Number of entities in the MOAlmanac database across cancer types. Bottom: Exact match accuracies from unstructured and structured RAG-LLM approaches across cancer types. **D.** Top: Number of entities in the MOAlmanac databas e across therapy strategies. Bottom: Exact match accuracies from unstructured and structured RAG-LLM approaches across therapy strategies. *Abbreviations in **C** and **D** are defined in Table S7*.

Additionally, structured data augmentation consistently outperformed the unstructured approach across various cancer types and therapy categories (Figure 3C–D). For example, an increase in accuracy of at least 30% was observed when transitioning from unstructured to structured augmentation for melanoma (MEL), prostate adenocarcinoma (PRAD), and cholangiocarcinoma (CHOL). Similarly, among therapy strategies, prompts related to antiandrogen + PARP inhibition, RAF inhibition, RAF + MEK inhibition, EGFR inhibition, and ER signaling inhibition also showed more than 30% improvement in accuracy with structured augmentation.

Of note, the model’s performance was suboptimal for certain therapies, particularly PARP and IDH1 inhibitors indicated for breast and ovarian cancer. This was primarily due to limitations in the retrieval step and the lack of reasoning to accurately link specific coding variants to their approved therapies, as well as difficulty distinguishing semantically granular contexts (e.g. treatment versus maintenance treatment).

However, the structured data augmented model performed strikingly well in some cases where structured clinicogenomic relationships were densely represented. Given that most antiandrogen + PARP inhibition therapies are approved for homologous recombination repair (HRR) gene mutations in prostate cancer—spanning 16 HRR genes linked to drug approvals— the high exact match accuracy achieved by the RAG-integrated model (LLM: 0%; unstructured RAG-LLM: 11%; structured RAG-LLM: 100%) suggests an effective linkage between HRR genes, approved therapies, and prostate cancer, which has the highest number of clinicogenomic associations in the dataset. Together, these results demonstrate that structured data empowers the model to better capture complex relationships between therapies and their approved indications, thereby improving performance across diverse clinical contexts.

### RAG-LLM accurately predicts therapies in real-world scenarios

To assess the real-world applicability of the RAG-LLM approach, we collected 21 clinical queries from 12 oncologists affiliated with Dana-Farber Cancer Institute. These queries focused on precision oncology therapies given a specific cancer type and biomarker(s). Of these, 10 queries had answers linked to on-label FDA-approved therapy indications listed in the MOAlmanac database (Table S5). We used the structured dataset as the context database for RAG and evaluated the model’s performance using exact match and partial match accuracies (Methods).

For queries where no on-label FDA-approved therapies currently exist, we evaluated whether RAG-LLM incorrectly retrieved off-label treatment options. We found that RAG-LLM exhibited lower performance on clinical queries (n=21) compared to synthetic queries (n=234), particularly exhibiting high rates of misattributions or hallucinations in cases without any appropriate FDA-approved drugs available (mean exact match accuracy: 50.48%; mean partial match accuracy: 65.71%; Figure 3A, 4A–B). For example, the LLM returned crizotinib, an FDA-approved ALK inhibitor for anaplastic large cell lymphoma, inflammatory myofibroblastic tumor, and non-small cell lung cancer, and misattributed it as an on-label therapy when queried about ALK inhibitor use for a patient with rhabdomyosarcoma and a *TFCP2* fusion (Supplementary Note 3).

However, when explicitly instructed to return no results for queries with no FDA-approved therapy available via an additional JSON schema (Methods), the model correctly returned no drugs in all such cases (Figure 4C), achieving a mean exact match accuracy of 80.95% and a mean partial match accuracy of 90.48%, significantly outperforming the version without the out-of-scope instruction (exact match: χ^2^(1) = 24.03, p = 1.90 x 10^-6^; partial match: χ^2^(1) = 13.59, p = 2.28 x 10^-4^; McNemar’s test on pooled results from five iterations; Figure 4A). Full outputs from the workflow are provided in Table S6. These results demonstrate the challenges in extrapolation of carefully designed queries to real-world queries, and potential strategies to counter common issues. They also demonstrate the effectiveness of RAG-LLM in retrieving relevant therapies for real-world oncologist queries, including those with no valid ground-truth answers.

**Figure 4.**
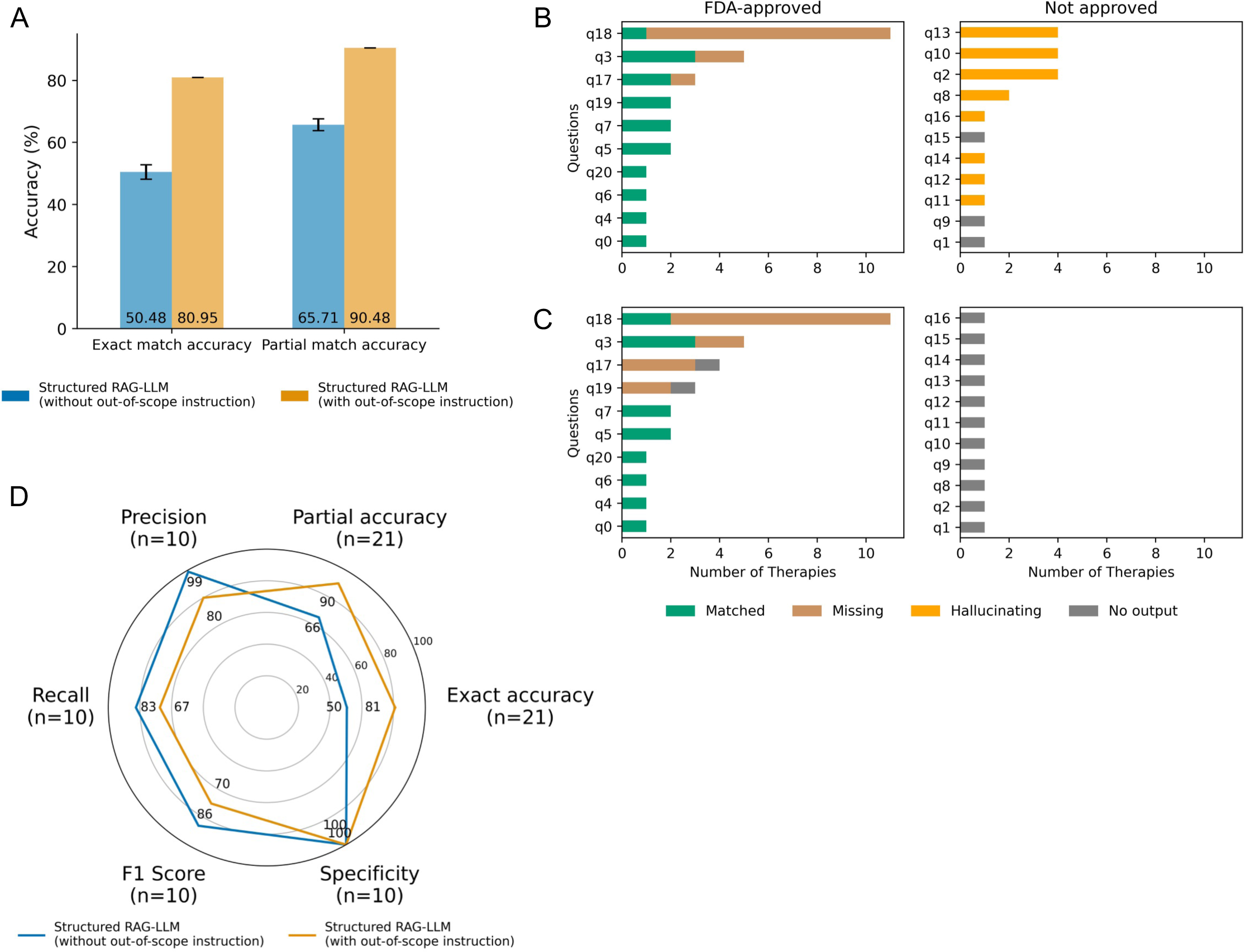
Performance of structured context-augmented RAG-LLM on real-world questions from practicing oncologists. **A.** Exact match and partial match accuracies from structured data-based RAG-LLM with or without explicit out-of-scope instructions. **B.** Number of matching, missing, hallucinated, and no-drug cases predicted by the workflow *without* explicit out-of-scope instructions, based on first-iteration predictions. Hallucinated drugs include all off-label drugs that were incorrectly attributed as FDA-approved. **C.** Same as **B**, but *with* explicit out-of-scope instructions (i.e., explicitly directing the model to return no matches via a JSON schema if no FDA-approved drugs are available). **D.** Average precision, recall, F1-score, specificity, and accuracy from structured data-augmented RAG-LLM. Accuracy was computed across all 21 queries; the remaining metrics were calculated across the 10 queries with FDA-approved therapies.

Although the out-of-scope instructed model achieved higher accuracy across all queries, the version without the out-of-scope instruction demonstrated better overall performance for the 10 questions with FDA-approved therapies (mean partial match accuracy: 100% vs. 80%; mean F1 score: 86% vs. 70%; Figure 4D). This version also achieved higher mean recall (83% vs. 67%) while preserving high mean precision (99% vs. 80%). In both strategies, average recall was slightly lower than precision. Notably, in at least six out of ten cases with FDA-approved therapies, the predicted therapies perfectly matched all the ground-truth therapies in both approaches. Together, these findings underscore the RAG-LLM approach’s effectiveness and strong reliability in making therapy predictions, while also highlighting how prompt design directly influences the model’s conservativeness—an aspect that can be flexibly tailored within our framework.

## Discussion

Broadly, this study investigated the potential of RAG-LLM to guide precision oncology decision-making across diverse use cases. We found that prompt optimization, particularly using a simple prompt, markedly improved LLM accuracy in retrieving FDA-approved biomarker-driven therapies, with structured data augmentation further boosting performance to achieve near-perfect accuracy in therapy predictions. Real-world oncologist queries validated the model’s ability to retrieve relevant therapies. Together, these findings highlight the transformative potential of RAG-LLM for precision oncology support.

Prompt design is essential for adapting general-purpose LLMs to specific applications^25^. We observed better performance with a basic prompt over a combined prompt, suggesting that enforcing JSON-format output may benefit from brevity. However, performance varied across models, especially smaller ones with fewer parameters, indicating no universal prompt design guarantees improvement across all LLMs in this use case.

RAG implementation significantly outperformed the LLM-only approach in predicting FDA-approved therapies, though performance varied with the format of the external dataset used. Our results demonstrate the model’s ability to extract relevant information from unstructured free text, while underscoring that highly curated structured data can significantly enhance its reliability, making the RAG-LLM built with structured data better suited for integration into clinical decision-making workflows.

We demonstrated the effectiveness of the RAG-LLM approach using a structured dataset in the real-world scenarios we evaluated but also identified an additional challenge. When no FDA-approved therapy was available for a given case, the model often misattributed treatments approved for other cancer types or biomarkers—although such treatments may still be used in practice based on their clinical relevance, this is beyond the scope of the current study. To address this issue, we implemented a predefined JSON schema instructing the model to return “no matches” when appropriate. This enhancement substantially improved accuracy with no misattributed cases. However, for queries with on-label FDA-approved drugs, the model without the out-of-scope instruction had better overall performance and higher recall while maintaining high precision compared to the out-of-scope instructed model, suggesting that strict adherence to the instruction may make the model more conservative in predicting drugs. In both versions, recall was lower than precision, indicating the framework is less likely to predict false positives, which is a critical feature in treatment recommendation tasks.

Ultimately, deploying LLM-based tools in clinical workflows demands strategies to mitigate the inherent risks arising from their widespread adoption. While the primary purpose of our RAG-LLM framework is to guide oncologists in recommending biomarker-driven therapies, its deployment will inevitably increase the risk of misuse or vulnerability to adversarial prompts and attacks—risks shared by all general-purpose LLMs^26^. Furthermore, integrating this framework into clinical settings will present unique challenges, including patient data privacy, potential harm from errors and biases, and protection of intellectual property and proprietary data^27^. Ensuring regulatory compliance with data privacy laws is also critical to maintaining patient confidentiality^28^. Thus, successful implementation of this framework is dependent on a comprehensive strategy that not only tackles the technical complexities of LLM deployment but also prospectively addresses the ethical and regulatory challenges in the high-stakes realm of oncology patient care.

In total, our RAG-LLM framework, designed to retrieve biomarker-based FDA-approved drugs, significantly outperformed an LLM-only implementation. Structured data augmentation further boosted its performance, and it demonstrated strong reliability when evaluated on real-world oncologist questions. Given that it requires fewer computational resources and greater adaptability than finetuning-based approaches, this framework may also facilitate greater equity in precision cancer medicine, particularly in supporting non-academic oncologists with limited resources. In addition, this flexible framework allows users to control the model’s conservativeness depending on their needs. For instance, for some rare cancers for which FDA-approvals are scarce, users can specify a higher temperature parameter and a less stringent prompt strategy to return a larger number of potentially relevant results, in addition to exact matches. Lastly, as the current process of staying up-to-date with regulatory approvals is highly fragmented, demanding toggling between multiple sources of information, our framework could also serve as a unified and reliable query layer across an otherwise fractured system.

### Limitations of the study

The primary focus of our study was to develop a reliable LLM-based framework that can serve as a decision-support tool for physicians in oncology. However, achieving the necessary accuracy and reliability for clinical deployment of this approach requires further refinements. Key areas for improvement include enhancing retrieval mechanisms to prioritize relevant therapies over purely text-matched results, expanding the context database by integrating clinical guidelines and clinical trial data to provide treatment options often available to physicians beyond FDA-approved drugs, and lastly, ensuring that treatment recommendations remain current by incorporating a dynamically updated knowledge base. Furthermore, increasing the number of real-world queries will improve the framework’s robustness by accounting for a more diverse range of actual patient cases. Addressing these enhancements will be essential for developing a clinically reliable, real-world applicable, and robust LLM-driven decision-support tool.

### Method details

The development and evaluation of this RAG-LLM precision oncology strategy involved three steps: 1) optimizing prompt design using a standard LLM, 2) evaluating the impact of both unstructured and structured context databases on the RAG-LLM’s ability to recommend biomarker-driven treatments, and 3) assessing the real-world applicability of the RAG-LLM by testing it with clinical queries from oncologists regarding biomarker-driven treatment recommendations.

### Database

To incorporate the latest knowledge on the clinical actionability of genomic biomarkers, we used FDA-approved drug indications from the April 11th, 2024 release of the Molecular Oncology Almanac (MOAlmanac) database (https://github.com/vanallenlab/moalmanac-db/releases/tag/v.2024-04-11). MOAlmanac contains both unstructured, free-text precision oncology genomic knowledge and structured data fields, including biomarkers, cancer types, and therapies^18^. Since the study’s primary use case is to assist medical professionals and molecular tumor boards in making treatment decisions, we focused exclusively on FDA-approved drugs to be conservative.

### Prompt engineering

To optimize prompt design for optimal model performance, we conducted a preliminary prompt engineering phase using the Mistral NeMo 12B model^29^, released in July 2024. Several prompt engineering strategies were evaluated, including:

1. Basic prompt: *“Please provide each line of treatment as a json format with the following JSON schema…Query: {prompt}”*
2. Scope-limiting prompt: *“Please only provide the therapies that are FDA-approved for the provided genomic biomarkers…Query: {prompt}”*
3. System role prompt: *“You are a helpful chatbot specialized in suggesting FDA-approved drugs to treat cancer…Query: {prompt}”*
4. Combination prompt: Merging strategies 2 and 3 with the basic prompt.

Additionally, the prompt included an example JSON schema to help the model generate structured output, which facilitated accurate and consistent evaluation (Table 1).

### Benchmark against other LLMs

In addition to the Mistral NeMo 12B model, we evaluated the performance of several other widely used LLMs. All models were accessed via their respective APIs, except Mistral 7B Instruct, an older model nearing deprecation, which was loaded and run locally on a Google Cloud virtual machine with 4 NVIDIA T4 GPUs.

– Mistral 7B Instruct (mistral-7B-Instruct-v0.3)

– Mistral 8B (ministral-8b-2410)

– Mistral 123B (mistral-large-2407)

– OpenAI GPT-4o (gpt-4o-2024-05-13)

– OpenAI GPT-4o-mini (gpt-4o-mini-2024-07-18)

– OpenAI o4-mini (o4-mini-2025-04-16)

### RAG-LLM approach

For the development and assessment of the RAG-LLM approach, we used 234 systematically generated prompts derived from each individual entity in the MOAlmanac database to mirror human-provided queries (Table S1–2). We used the top-ranked GPT-4o model for all subsequent analyses.

To evaluate the RAG-LLM approach using an unstructured dataset, we generated a dataset of FDA drug label text in PDF format for each biomarker-based therapy approval and extracted each label’s ‘Indication and Usage’ section. We selected drug labels for FDA-approved oncology therapies involving a biomarker for at least one approved indication, as curated by the MOAlmanac database. The resulting unstructured dataset consisted of 56 original ‘Indications and Usage’ sections, which served as the context database for RAG integration (Table S3). A representative input-output example using unstructured data is available in Supplementary Note 1.

To evaluate the RAG-LLM approach using a structured dataset, we manually created an answer set for the synthetic prompts, incorporating it into the RAG-LLM workflow as a structured context database. Each answer chunk corresponded to each entity or therapy-biomarker relationship in MOAlamanc (Table S4). An example of an input prompt augmented with structured data and its output from the RAG-LLM workflow is provided in Supplementary Note 2. For both approaches, the context database was embedded using the *text-embedding-3-small* model^30^ and stored as a vector database via the *FAISS* library^31^. Each query embedding was then compared to the embedded context database using Euclidean distance, retrieving the top 10 most similar text chunks to supplement the prompt fed into the LLM for inference.

### Real-world question survey and evaluation

To collect real-world clinical questions, we designed a survey and distributed it to collaborating physicians at the Dana-Farber Cancer Institute and Boston Children’s Hospital. The survey introduced the MOAlmanac database, focusing on FDA-approved drug indications and clinical genomic biomarkers. Physicians were asked to submit questions related to their clinical practice regarding drug actionability, treatment regimens, or biomarker associations, without including identifiable patient data. These responses were then used as prompts to evaluate our RAG-LLM approach, assessing its ability to generate relevant and accurate answers in real-world clinical settings. The prompt design was refined to handle cases where no FDA-approved therapies exist. To account for variability across model runs, we conducted five iterations of the RAG-LLM workflow.

### Evaluation

We evaluated LLM performance with and without RAG by calculating the proportion of exact and partial matches of correctly predicted therapy recommendations across all the relationships. Each LLM output was generated in a JSON format and was parsed line by line for drug names following the ‘Drug Name’ entity to compute the accuracy. To account for the non-deterministic behavior of LLMs, we ran five iterations of the workflow and averaged the accuracy across iterations. For exact match accuracy, predictions were considered correct if all the ground-truth therapies (Table S2) were present in the drug output. For partial match accuracy, predictions were considered correct if at least one ground truth therapy was present:

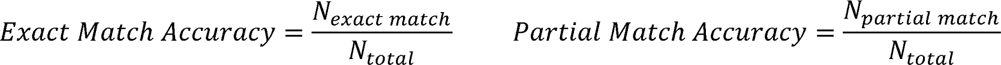

Where:

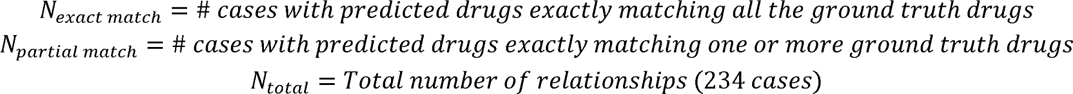

Additionally, we calculated precision, recall, F1 score, and specificity to comprehensively assess model performance across all queries. To minimize the randomness in outputs, we set the temperature to 0.0 and initialized a fixed random seed.

### Statistical test

We performed McNemar’s test to compare accuracies between different approaches. When the total number of discordant pairs was 25 or greater, we used the chi-square approximation with a continuity correction was used to prevent overestimation of significance. For fewer than 25 discordant pairs, we applied the exact binomial test. To assess overall performance across iterations, we pooled the discordant pair counts from all runs and then performed the McNemar’s test. When comparing evaluation metrics other than accuracy, we used a one-sided Wilcoxon signed-rank test. To account for multiple hypothesis testing, p-values were adjusted using the Benjamini-Hochberg correction.

## Resource availability

### Data and code availability

All the scripts and datasets for running the LLM and RAG-LLM pipelines, along with the corresponding outputs generated in this study, are publicly available at https://github.com/hjjshine/rag-llm-cancer-paper. Pipeline usage instructions are provided in the README file. Model’s conservativeness can be controlled using the temperature and strategy parameters.

## Data Availability

All data produced are available online at https://github.com/hjjshine/rag-llm-cancer-paper

## Acknowledgements

This work was supported by P50CA272390, DOD W81XWH-21-PCRP-DSA, DOD HT94252410415, and the Mark Foundation Emerging Leader Award.

