## Supplementary Appendix 1 for "Implementing a context-augmented large language model to guide precision cancer medicine"

**Supplementary Note**

Supplementary Note 1. Example of a paired prompt-output from RAG-LLM with unstructured data

**Prompt:**

Context information is below.

---------------------

['INDICATIONS AND USAGE -------------------------- \nLynparza is a poly (ADP-ribose) polymerase (PARP) inhibitor indicated: \nOvarian cancer \n• for the maintenance treatment of adult patients with deleterious or \nsuspected deleterious germline or somatic BRCA-mutated advanced \nepithelial ovarian, fallopian tube or primary peritoneal cancer who are in \ncomplete or partial response to first-line platinum-based chemotherapy. \nSelect patients for therapy based on an FDA-approved companion \ndiagnostic for Lynparza. (1.1, 2.1) \n• in combination with bevacizumab for the maintenance treatment of adult \npatients with advanced epithelial ovarian, fallopian tube or primary \nperitoneal cancer who are in complete or partial response to first-line \nplatinum-based chemotherapy and whose cancer is associated with \nhomologous recombination deficiency (HRD)-positive status defined by \neither: \n• a deleterious or suspected deleterious BRCA mutation, and/or \n• genomic instability. \nSelect patients for therapy based on an FDA-approved companion \ndiagnostic for Lynparza (1.2, 2.1). \n• for the maintenance treatment of adult patients with recurrent epithelial \novarian, fallopian tube or primary peritoneal cancer, who are in \ncomplete or partial response to platinum-based chemotherapy. (1.3) \nBreast cancer \n• for the adjuvant treatment of adult patients with deleterious or suspected \ndeleterious gBRCAm human epidermal growth factor receptor 2 \n(HER2)-negative high risk early breast cancer who have been treated \nwith neoadjuvant or adjuvant chemotherapy. Select patients for therapy \nbased on an FDA-approved companion diagnostic for Lynparza. (1.4, \n2.1). \n• for the treatment of adult patients with deleterious or suspected \ndeleterious gBRCAm, HER2-negative metastatic breast cancer who have \nbeen treated with chemotherapy in the neoadjuvant, adjuvant or \nmetastatic setting. Patients with hormone receptor (HR)-positive breast \ncancer should have been treated with a prior endocrine therapy or be \nconsidered inappropriate for endocrine therapy. Select patients for \ntherapy based on an FDA-approved companion diagnostic for Lynparza. \n(1.5, 2.1) \nPancreatic cancer \n• for the maintenance treatment of adult patients with deleterious or \nsuspected deleterious gBRCAm metastatic pancreatic adenocarcinoma \nwhose disease has not progressed on at least 16 weeks of a first-line \nplatinum-based chemotherapy regimen. Select patients for therapy based \non an FDA-approved companion diagnostic for Lynparza. (1.6, 2.1) \nProstate cancer \n• for the treatment of adult patients with deleterious or suspected \ndeleterious germline or somatic homologous recombination repair \n(HRR) gene-mutated metastatic castration-resistant prostate cancer \n(mCRPC) who have progressed following prior treatment with \nenzalutamide or abiraterone. Select patients for therapy based on an \nFDA-approved companion diagnostic for Lynparza. (1.7, 2.1) \n• in combination with abiraterone and prednisone or prednisolone for the \ntreatment of adult patients with deleterious or suspected deleterious \nBRCA-mutated (BRCAm) metastatic castration-resistant prostate cancer \n(mCRPC). Select patients for therapy based on an FDA-approved \ncompanion diagnostic for Lynparza. (1.8, 2.1) \n ----------------------', 'INDICATIONS AND USAGE---------------------------\xad\nRUBRACA is a poly (ADP-ribose) polymerase (PARP) inhibitor indicated: \nOvarian cancer \n•\t for the maintenance treatment of adult patients with recurrent epithelial \novarian, fallopian tube, or primary peritoneal cancer who are in a complete \nor partial response to platinum-based chemotherapy. (1.1) \n•\t for the treatment of adult patients with a deleterious BRCA mutation \n(germline and/or somatic)-associated epithelial ovarian, fallopian tube, or \nprimary peritoneal cancer who have been treated with two or more \nchemotherapies. Select patients for therapy based on an FDA-approved \ncompanion diagnostic for RUBRACA. (1.1, 2.1) \nProstate cancer \n•\t for the treatment of adult patients with a deleterious BRCA mutation \n(germline and/or somatic)-associated metastatic castration-resistant prostate \ncancer (mCRPC) who have been treated with androgen receptor-directed \ntherapy and a taxane-based chemotherapy. Select patients for therapy based \non an FDA-approved companion diagnostic for RUBRACA. (1.2, 2.1) \nThis indication is approved under accelerated approval based on objective \nresponse rate and duration of response. Continued approval for this \nindication may be contingent upon verification and description of clinical \nbenefit in confirmatory trials. (1.2) \n-----------------------', 'INDICATIONS AND USAGE----------------------------\nKEYTRUDA is a programmed death receptor-1 (PD-1)-blocking \nantibody indicated: \nMelanoma \n• for the treatment of patients with unresectable or metastatic \nmelanoma. (1.1) \n• for the adjuvant treatment of adult and pediatric (12 years and \nolder) patients with Stage IIB, IIC, or III melanoma following \ncomplete resection. (1.1) \nNon-Small Cell Lung Cancer (NSCLC) \n• in combination with pemetrexed and platinum chemotherapy, \nas first-line treatment of patients with metastatic nonsquamous \nNSCLC, with no EGFR or ALK genomic tumor aberrations. \n(1.2) \n• in combination with carboplatin and either paclitaxel or \npaclitaxel protein-bound, as first-line treatment of patients with \nmetastatic squamous NSCLC. (1.2) \n• as a single agent for the first-line treatment of patients with \nNSCLC expressing PD-L1 [Tumor Proportion Score (TPS) \n≥1%] as determined by an FDA-approved test, with no EGFR \nor ALK genomic tumor aberrations, and is: \no Stage III where patients are not candidates for surgical \nresection or definitive chemoradiation, or \no metastatic. (1.2, 2.1) \n• as a single agent for the treatment of patients with metastatic \nNSCLC whose tumors express PD-L1 (TPS ≥1%) as \ndetermined by an FDA-approved test, with disease \nprogression on or after platinum-containing chemotherapy. \nPatients with EGFR or ALK genomic tumor aberrations should \nhave disease progression on FDA-approved therapy for these \naberrations prior to receiving KEYTRUDA. (1.2, 2.1) \n• for the treatment of patients with resectable (tumors ≥4 cm or \nnode positive) NSCLC in combination with platinum-containing \nchemotherapy as neoadjuvant treatment, and then continued \nas a single agent as adjuvant treatment after surgery. (1.2) \n• as a single agent, for adjuvant treatment following resection \nand platinum-based chemotherapy for adult patients with \nStage IB (T2a ≥4 cm), II, or IIIA NSCLC. (1.2) \nHead and Neck Squamous Cell Cancer (HNSCC) \n• in combination with platinum and FU for the first-line treatment \nof patients with metastatic or with unresectable, recurrent \nHNSCC. (1.3) \n• as a single agent for the first-line treatment of patients with \nmetastatic or with unresectable, recurrent HNSCC whose \ntumors express PD-L1 [Combined Positive Score (CPS) ≥1] as \ndetermined by an FDA-approved test. (1.3, 2.1) \n• as a single agent for the treatment of patients with recurrent or \nmetastatic HNSCC with disease progression on or after \nplatinum-containing chemotherapy. (1.3) \nClassical Hodgkin Lymphoma (cHL) \n• for the treatment of adult patients with relapsed or refractory \ncHL. (1.4) \n• for the treatment of pediatric patients with refractory cHL, or \ncHL that has relapsed after 2 or more lines of therapy. (1.4) \nPrimary Mediastinal Large B-Cell Lymphoma (PMBCL) \n• for the treatment of adult and pediatric patients with refractory \nPMBCL, or who have relapsed after 2 or more prior lines of \ntherapy. (1.5) \n• Limitations of Use: KEYTRUDA is not recommended for \ntreatment of patients with PMBCL who require urgent \ncytoreductive therapy. \nUrothelial Carcinoma \n• in combination with enfortumab vedotin, for the treatment of \nadult patients with locally advanced or metastatic urothelial \ncarcinoma who are not eligible for cisplatin-containing \nchemotherapy.1 (1.6) \n• as a single agent for the treatment of patients with locally \nadvanced or metastatic urothelial carcinoma who: \no are not eligible for any platinum-containing \nchemotherapy, or \no who have disease progression during or following \nplatinum-containing chemotherapy or within 12 months of \nneoadjuvant or adjuvant treatment with platinum-\ncontaining chemotherapy. (1.6) \n• as a single agent for the treatment of patients with Bacillus \nCalmette-Guerin (BCG)-unresponsive, high-risk, non-muscle \ninvasive bladder cancer (NMIBC) with carcinoma in situ (CIS) \nwith or without papillary tumors who are ineligible for or have \nelected not to undergo cystectomy. (1.6) \nMicrosatellite Instability-High or Mismatch Repair Deficient Cancer \n• for the treatment of adult and pediatric patients with \nunresectable or metastatic microsatellite instability-high \n(MSI-H) or mismatch repair deficient (dMMR) solid tumors, as \ndetermined by an FDA-approved test, that have progressed \nfollowing prior treatment and who have no satisfactory \nalternative treatment options. (1.7, 2.1) \nMicrosatellite Instability-High or Mismatch Repair Deficient \nColorectal Cancer (CRC) \n• for the treatment of patients with unresectable or metastatic \nMSI-H or dMMR colorectal cancer (CRC) as determined by an \nFDA-approved test. (1.8, 2.1) \nGastric Cancer \n• in combination with trastuzumab, fluoropyrimidine- and \nplatinum-containing chemotherapy, for the first-line treatment \nof patients with locally advanced unresectable or metastatic \nHER2-positive gastric or gastroesophageal junction (GEJ) \nadenocarcinoma whose tumors express PD-L1 (CPS ≥1) as \ndetermined by an FDA-approved test.1 (1.9) \nEsophageal Cancer \n• for the treatment of patients with locally advanced or metastatic \nesophageal or gastroesophageal junction (GEJ) (tumors with \nepicenter 1 to 5 centimeters above the GEJ) carcinoma that is \nnot amenable to surgical resection or definitive chemoradiation \neither: \no in combination with platinum- and fluoropyrimidine-based \nchemotherapy, or \no as a single agent after one or more prior lines of systemic \ntherapy for patients with tumors of squamous cell \nhistology that express PD-L1 (CPS ≥10) as determined \nby an FDA-approved test. (1.10, 2.1) \nCervical Cancer \n• in combination with chemotherapy, with or without \nbevacizumab, for the treatment of patients with persistent, \nrecurrent, or metastatic cervical cancer whose tumors express \nPD-L1 (CPS ≥1) as determined by an FDA-approved test. \n(1.11, 2.1) \n• as a single agent for the treatment of patients with recurrent or \nmetastatic cervical cancer with disease progression on or after \nchemotherapy whose tumors express PD-L1 (CPS ≥1) as \ndetermined by an FDA-approved test. (1.11, 2.1) \nHepatocellular Carcinoma (HCC) \n• for the treatment of patients with HCC who have been \npreviously treated with sorafenib.1 (1.12) \nBiliary Tract Cancer (BTC) \n• in combination with gemcitabine and cisplatin, for the \ntreatment of patients with locally advanced unresectable or \nmetastatic biliary tract cancer. (1.13) \nMerkel Cell Carcinoma (MCC) \n• for the treatment of adult and pediatric patients with recurrent \nlocally advanced or metastatic Merkel cell carcinoma. (1.14) \nRenal Cell Carcinoma (RCC) \n• in combination with axitinib, for the first-line treatment of adult \npatients with advanced RCC. (1.15) \n• in combination with lenvatinib, for the first-line treatment of \nadult patients with advanced RCC. (1.15) \n• for the adjuvant treatment of patients with RCC at \nintermediate-high or high risk of recurrence following \nReference ID: 5274826 \nThis label may not be the latest approved by FDA. \nFor current labeling information, please visit https://www.fda.gov/drugsatfda \nnephrectomy, or following nephrectomy and resection of \nmetastatic lesions. (1.15) \nEndometrial Carcinoma \n• in combination with lenvatinib, for the treatment of patients with \nadvanced endometrial carcinoma that is mismatch repair \nproficient (pMMR) as determined by an FDA-approved test or \nnot MSI-H, who have disease progression following prior \nsystemic therapy in any setting and are not candidates for \ncurative surgery or radiation. (1.16, 2.1) \n• as a single agent, for the treatment of patients with advanced \nendometrial carcinoma that is MSI-H or dMMR, as determined \nby an FDA-approved test, who have disease progression \nfollowing prior systemic therapy in any setting and are not \ncandidates for curative surgery or radiation. (1.16, 2.1) \nTumor Mutational Burden-High (TMB-H) Cancer \n• for the treatment of adult and pediatric patients with \nunresectable or metastatic tumor mutational burden-high \n(TMB-H) [≥10 mutations/megabase (mut/Mb)] solid tumors, as \ndetermined by an FDA-approved test, that have progressed \nfollowing prior treatment and who have no satisfactory \nalternative treatment options.1 (1.17, 2.1) \n• Limitations of Use: The safety and effectiveness of \nKEYTRUDA in pediatric patients with TMB-H central nervous \nsystem cancers have not been established. \nCutaneous Squamous Cell Carcinoma (cSCC) \n• for the treatment of patients with recurrent or metastatic cSCC \nor locally advanced cSCC that is not curable by surgery or \nradiation. (1.18) \nTriple-Negative Breast Cancer (TNBC) \n• for the treatment of patients with high-risk early-stage TNBC in \ncombination with chemotherapy as neoadjuvant treatment, and \nthen continued as a single agent as adjuvant treatment after \nsurgery. (1.19) \n• in combination with chemotherapy, for the treatment of \npatients with locally recurrent unresectable or metastatic \nTNBC whose tumors express PD-L1 (CPS ≥10) as determined \nby an FDA approved test. (1.19, 2.1) \nAdult Classical Hodgkin Lymphoma and Adult Primary Mediastinal \nLarge B-Cell Lymphoma: Additional Dosing Regimen of 400 mg \nEvery 6 Weeks \n• for use at an additional recommended dosage of 400 mg every \n6 weeks for Classical Hodgkin Lymphoma and Primary \nMediastinal Large B-Cell Lymphoma in adults.2 (1.20, 2.2) \n1 This indication is approved under accelerated approval based on \ntumor response rate and durability of response. Continued \napproval for this indication may be contingent upon verification \nand description of clinical benefit in the confirmatory trials. \n2 This indication is approved under accelerated approval based on \npharmacokinetic data, the relationship of exposure to efficacy, \nand the relationship of exposure to safety. Continued approval for \nthis dosing may be contingent upon verification and description of \nclinical benefit in the confirmatory trials. \n-----------------------', 'INDICATIONS AND USAGE---------------------------\nAKEEGA is a combination of niraparib, a poly (ADP-ribose) polymerase \n(PARP) inhibitor, and abiraterone acetate, a CYP17 inhibitor indicated with \nprednisone for the treatment of adult patients with deleterious or suspected \ndeleterious BRCA-mutated (BRCAm) metastatic castration-resistant prostate \ncancer (mCRPC). Select patients for therapy based on an FDA-approved test \nfor AKEEGA. (1, 2.1) \n------------------------', 'INDICATIONS AND USAGE----------------------------\nTALZENNA is a poly (ADP-ribose) polymerase (PARP) inhibitor indicated \nfor: \nBreast Cancer \n• As a single agent, for the treatment of adult patients with deleterious or \nsuspected deleterious germline BRCA-mutated (gBRCAm) HER2-negative \nlocally advanced or metastatic breast cancer. Select patients for therapy \nbased on an FDA-approved companion diagnostic for TALZENNA. (1.1) \nHRR Gene-mutated mCRPC \n• In combination with enzalutamide for the treatment of adult patients with \nHRR gene-mutated metastatic castration-resistant prostate cancer \n(mCRPC). (1.2) \n----------------------', 'INDICATIONS AND USAGE---------------------------\nBALVERSA is a kinase inhibitor indicated for the treatment of adult patients \nwith locally advanced or metastatic urothelial carcinoma (mUC) with \nsusceptible FGFR3 genetic alterations whose disease has progressed on or \nafter at least one line of prior systemic therapy. \nSelect patients for therapy based on an FDA-approved companion diagnostic \nfor BALVERSA. (1, 2.1) \nLimitations of Use \nBALVERSA is not recommended for the treatment of patients who are \neligible for and have not received prior PD-1 or PD-L1 inhibitor therapy. (1, \n14.1) \n------------------------', 'INDICATIONS AND USAGE-------------------------\nBRAFTOVI is a kinase inhibitor indicated: \n\uf0b7 in combination with binimetinib, for the treatment of patients with \nunresectable or metastatic melanoma with a BRAF V600E or V600K \nmutation, as detected by an FDA-approved test. (1.1, 2.1)\n\uf0b7 in combination with cetuximab, for the treatment of adult patients \nwith metastatic colorectal cancer (CRC) with a BRAF V600E \nmutation, as detected by an FDA-approved test, after prior therapy. \n(1.2, 2.1)\n\uf0b7 in combination with binimetinib, for the treatment of adult patients \nwith metastatic non-small cell lung cancer (NSCLC) with a BRAF \nV600E mutation, as detected by an FDA-approved test. (1.3, 2.1)\nLimitations of Use\nBRAFTOVI is not indicated for treatment of patients with wild-type BRAF \nmelanoma, wild-type BRAF CRC, or wild-type BRAF NSCLC. (1.4, 5.2)\n---------------------', 'INDICATIONS AND USAGE------------------------------ \nTYKERB is a kinase inhibitor indicated in combination with: (1) \n\uf0b7 capecitabine for the treatment of patients with advanced or metastatic breast \ncancer whose tumors overexpress human epidermal growth factor receptor 2 \n(HER2) and who have received prior therapy including an anthracycline, a \ntaxane, and trastuzumab. \nLimitations of Use: Patients should have disease progression on trastuzumab \nprior to initiation of treatment with TYKERB in combination with capecitabine. \n\uf0b7 letrozole for the treatment of postmenopausal women with hormone receptor-\npositive metastatic breast cancer that overexpresses the HER2 receptor for \nwhom hormonal therapy is indicated. \nTYKERB in combination with an aromatase inhibitor has not been compared to a \ntrastuzumab-containing chemotherapy regimen for the treatment of metastatic \nbreast cancer. \n----------------------------', 'INDICATIONS AND USAGE---------------------------\nRYBREVANT is a bispecific EGF receptor-directed and MET receptor-\ndirected antibody indicated: \n• in combination with carboplatin and pemetrexed for the first-line treatment \nof adult patients with locally advanced or metastatic non-small cell lung cancer \n(NSCLC) with epidermal growth factor receptor (EGFR) exon 20 insertion \nmutations, as detected by an FDA-approved test. (1, 2.2) \n• as a single agent for the treatment of adult patients with locally advanced or \nmetastatic NSCLC with EGFR exon 20 insertion mutations, as detected by an \nFDA-approved test, whose disease has progressed on or after platinum-based \nchemotherapy. (1, 2.2) \n------------------------', 'INDICATIONS AND USAGE -------------------------\xad\nSTIVARGA is a kinase inhibitor indicated for the treatment of patients with: \n• Metastatic colorectal cancer (CRC) who have been previously treated with \nfluoropyrimidine-, oxaliplatin- and irinotecan-based chemotherapy, an anti-\nVEGF therapy, and, if RAS wild-type, an anti-EGFR therapy. (1.1) \n• Locally advanced, unresectable or metastatic gastrointestinal stromal tumor \n(GIST) who have been previously treated with imatinib mesylate and \nsunitinib malate. (1.2) \n• Hepatocellular carcinoma (HCC) who have been previously treated with \nsorafenib (1.3) \n----------------------']

---------------------

Please provide each treatment as a json format with the following JSON schema.

{

"Treatment 1": {

"Disease Name": ,

"Disease Phase or Condition": ,

"Drug Name": ,

"Prior Treatment or Resistance Status": ,

"Genomic Features":

"Link to FDA-approved Label":

}

}

Query: If a prostate cancer patient has a somatic variant in gene BRCA1, and is metastatic castration-resistant, what would be the suggested lines of treatment?

**Output:**

{

"Treatment 1": {

"Disease Name": "Prostate Cancer",

"Disease Phase or Condition": "Metastatic Castration-Resistant",

"Drug Name": "Lynparza",

"Prior Treatment or Resistance Status": "Progressed following prior treatment with enzalutamide or abiraterone",

"Genomic Features": "Deleterious or suspected deleterious germline or somatic homologous recombination repair (HRR) gene-mutated",

"Link to FDA-approved Label": "https://www.fda.gov/drugsatfda"

},

"Treatment 2": {

"Disease Name": "Prostate Cancer",

"Disease Phase or Condition": "Metastatic Castration-Resistant",

"Drug Name": "Lynparza in combination with abiraterone and prednisone or prednisolone",

"Prior Treatment or Resistance Status": "None specified",

"Genomic Features": "Deleterious or suspected deleterious BRCA-mutated (BRCAm)",

"Link to FDA-approved Label": "https://www.fda.gov/drugsatfda"

},

"Treatment 3": {

"Disease Name": "Prostate Cancer",

"Disease Phase or Condition": "Metastatic Castration-Resistant",

"Drug Name": "RUBRACA",

"Prior Treatment or Resistance Status": "Treated with androgen receptor-directed therapy and a taxane-based chemotherapy",

"Genomic Features": "Deleterious BRCA mutation (germline and/or somatic)",

"Link to FDA-approved Label": "https://www.fda.gov/drugsatfda"

},

"Treatment 4": {

"Disease Name": "Prostate Cancer",

"Disease Phase or Condition": "Metastatic Castration-Resistant",

"Drug Name": "AKEEGA (niraparib and abiraterone acetate) with prednisone",

"Prior Treatment or Resistance Status": "None specified",

"Genomic Features": "Deleterious or suspected deleterious BRCA-mutated (BRCAm)",

"Link to FDA-approved Label": "https://www.fda.gov/drugsatfda"

},

"Treatment 5": {

"Disease Name": "Prostate Cancer",

"Disease Phase or Condition": "Metastatic Castration-Resistant",

"Drug Name": "TALZENNA in combination with enzalutamide",

"Prior Treatment or Resistance Status": "None specified",

"Genomic Features": "HRR gene-mutated",

"Link to FDA-approved Label": "https://www.fda.gov/drugsatfda"

}

}

Supplementary Note 2. Example of a paired prompt-output from RAG-LLM with structured data

**Prompt:**

Context information is below.

---------------------

['If a prostate cancer patient has a somatic variant in gene BRCA1, and is metastatic castration-resistant, one recommended treatment would be Abiraterone + Prednisone + Olaparib. The U.S. Food and Drug Administration (FDA) has granted approval to olaparib in combination with abiraterone and prednisone or prednisolone for the treatment of adult patients with deleterious or suspected deleterious BRCA-mutated (BRCAm) metastatic castration-resistant prostate cancer (mCRPC). (Citation: AstraZeneca Pharmaceuticals, LP. Lynparza (olaparib) [package insert]. U.S. Food and Drug Administration website. https://www.accessdata.fda.gov/drugsatfda_docs/label/2023/208558s025lbl.pdf. Revised May 2023. Accessed July 5, 2023.).', 'If a prostate cancer patient has a somatic variant in gene BRCA2, and is metastatic castration-resistant, one recommended treatment would be Abiraterone + Prednisone + Olaparib. The U.S. Food and Drug Administration (FDA) has granted approval to olaparib in combination with abiraterone and prednisone or prednisolone for the treatment of adult patients with deleterious or suspected deleterious BRCA-mutated (BRCAm) metastatic castration-resistant prostate cancer (mCRPC). (Citation: AstraZeneca Pharmaceuticals, LP. Lynparza (olaparib) [package insert]. U.S. Food and Drug Administration website. https://www.accessdata.fda.gov/drugsatfda_docs/label/2023/208558s025lbl.pdf. Revised May 2023. Accessed July 5, 2023.).', 'If a prostate cancer patient has a somatic variant in gene BRCA1, and is metastatic castration-resistant, one recommended treatment would be Enzalutamide + Talazoparib. The U.S. Food and Drug Administration (FDA) granted approval to talazoparib in combination with enzalutamide for the treatment of adult patients with HRR gene-mutated metastatic castration-resistant prostate cancer (mCRPC). (Citation: Pfizer, Inc. Talzenna (talazoparib) [package insert]. U.S. Food and Drug Administration website. https://www.accessdata.fda.gov/drugsatfda_docs/label/2023/211651s010lbl.pdf. Revised June 2023. Accessed July 6, 2023.).', 'If a prostate cancer patient has a somatic variant in gene BRCA1, and is metastatic castration resistant prostate cancer, one recommended treatment would be Olaparib. The U.S. Food and Drug Administration (FDA) granted approval to olaparib for adult patients with deleterious or suspected deleterous germline or somatic homologous recombination repair (HRR) gene-mutated metastatic castration-resistant prostate cancer (mCRPC), who have progressed following prior treatment with enzalutamide or abiraterone. (Citation: AstraZeneca Pharmaceuticals, LP. Lynparza (olaparib) [package insert]. U.S. Food and Drug Administration website. https://www.accessdata.fda.gov/drugsatfda_docs/label/2023/208558s025lbl.pdf. Revised May 2023. Accessed July 5, 2023.).', 'If a prostate cancer patient has a somatic variant in gene BRCA2, and is metastatic castration-resistant, one recommended treatment would be Enzalutamide + Talazoparib. The U.S. Food and Drug Administration (FDA) granted approval to talazoparib in combination with enzalutamide for the treatment of adult patients with HRR gene-mutated metastatic castration-resistant prostate cancer (mCRPC). (Citation: Pfizer, Inc. Talzenna (talazoparib) [package insert]. U.S. Food and Drug Administration website. https://www.accessdata.fda.gov/drugsatfda_docs/label/2023/211651s010lbl.pdf. Revised June 2023. Accessed July 6, 2023.).', 'If a prostate cancer patient has a somatic variant in gene BRCA2, and is metastatic castration resistant prostate cancer, one recommended treatment would be Olaparib. The U.S. Food and Drug Administration (FDA) granted approval to olaparib for adult patients with deleterious or suspected deleterous germline or somatic homologous recombination repair (HRR) gene-mutated metastatic castration-resistant prostate cancer (mCRPC), who have progressed following prior treatment with enzalutamide or abiraterone. (Citation: AstraZeneca Pharmaceuticals, LP. Lynparza (olaparib) [package insert]. U.S. Food and Drug Administration website. https://www.accessdata.fda.gov/drugsatfda_docs/label/2023/208558s025lbl.pdf. Revised May 2023. Accessed July 5, 2023.).', 'If a prostate cancer patient has a somatic variant in gene BRIP1, and is metastatic castration resistant prostate cancer, one recommended treatment would be Olaparib. The U.S. Food and Drug Administration (FDA) granted approval to olaparib for adult patients with deleterious or suspected deleterous germline or somatic homologous recombination repair (HRR) gene-mutated metastatic castration-resistant prostate cancer (mCRPC), who have progressed following prior treatment with enzalutamide or abiraterone. (Citation: AstraZeneca Pharmaceuticals, LP. Lynparza (olaparib) [package insert]. U.S. Food and Drug Administration website. https://www.accessdata.fda.gov/drugsatfda_docs/label/2023/208558s025lbl.pdf. Revised May 2023. Accessed July 5, 2023.).', 'If a prostate cancer patient has a germline variant in gene BRCA1, and is metastatic castration-resistant, one recommended treatment would be Abiraterone + Prednisone + Olaparib. The U.S. Food and Drug Administration (FDA) has granted approval to olaparib in combination with abiraterone and prednisone or prednisolone for the treatment of adult patients with deleterious or suspected deleterious BRCA-mutated (BRCAm) metastatic castration-resistant prostate cancer (mCRPC). (Citation: AstraZeneca Pharmaceuticals, LP. Lynparza (olaparib) [package insert]. U.S. Food and Drug Administration website. https://www.accessdata.fda.gov/drugsatfda_docs/label/2023/208558s025lbl.pdf. Revised May 2023. Accessed July 5, 2023.).', 'If a prostate cancer patient has a somatic variant in gene BRCA1, and is metastatic castration-resistant, one recommended treatment would be Abiraterone acetate + Niraparib. The U.S. Food and Drug Administration (FDA) granted approval to niraparib, a poly (ADP-ribose) polymerase (PARP) inhibitor, and abiraterone acetate, a CYP17 inhibitor indicated with prednisone for the treatment of adult patients with deleterious or suspected deleterious BRCA-mutated (BRCAm) metastatic castration-resistant prostate cancer (mCRPC). (Citation: Janssen Biotech, Inc. Akeega (niraparib and abiraterone acetate) [package insert]. U.S. Food and Drug Administration website. https://www.accessdata.fda.gov/drugsatfda_docs/label/2023/216793s000lbl.pdf. Revised August 2023. Accessed September 6, 2023.).', 'If a prostate cancer patient has a somatic variant in gene BRCA2, and is metastatic castration-resistant, one recommended treatment would be Abiraterone acetate + Niraparib. The U.S. Food and Drug Administration (FDA) granted approval to niraparib, a poly (ADP-ribose) polymerase (PARP) inhibitor, and abiraterone acetate, a CYP17 inhibitor indicated with prednisone for the treatment of adult patients with deleterious or suspected deleterious BRCA-mutated (BRCAm) metastatic castration-resistant prostate cancer (mCRPC). (Citation: Janssen Biotech, Inc. Akeega (niraparib and abiraterone acetate) [package insert]. U.S. Food and Drug Administration website. https://www.accessdata.fda.gov/drugsatfda_docs/label/2023/216793s000lbl.pdf. Revised August 2023. Accessed September 6, 2023.).']

---------------------

Please provide each treatment as a json format with the following JSON schema.

{

"Treatment 1": {

"Disease Name": ,

"Disease Phase or Condition": ,

"Drug Name": ,

"Prior Treatment or Resistance Status": ,

"Genomic Features":

"Link to FDA-approved Label":

}

}

Query: If a prostate cancer patient has a somatic variant in gene BRCA1, and is metastatic castration-resistant, what would be the suggested lines of treatment?

**Output:**

{

"Treatment 1": {

"Disease Name": "Prostate Cancer",

"Disease Phase or Condition": "Metastatic Castration-Resistant",

"Drug Name": "Abiraterone + Prednisone + Olaparib",

"Prior Treatment or Resistance Status": "Not specified",

"Genomic Features": "Somatic variant in gene BRCA1",

"Link to FDA-approved Label": "https://www.accessdata.fda.gov/drugsatfda_docs/label/2023/208558s025lbl.pdf"

},

"Treatment 2": {

"Disease Name": "Prostate Cancer",

"Disease Phase or Condition": "Metastatic Castration-Resistant",

"Drug Name": "Enzalutamide + Talazoparib",

"Prior Treatment or Resistance Status": "Not specified",

"Genomic Features": "Somatic variant in gene BRCA1",

"Link to FDA-approved Label": "https://www.accessdata.fda.gov/drugsatfda_docs/label/2023/211651s010lbl.pdf"

},

"Treatment 3": {

"Disease Name": "Prostate Cancer",

"Disease Phase or Condition": "Metastatic Castration-Resistant",

"Drug Name": "Olaparib",

"Prior Treatment or Resistance Status": "Progressed following prior treatment with enzalutamide or abiraterone",

"Genomic Features": "Somatic variant in gene BRCA1",

"Link to FDA-approved Label": "https://www.accessdata.fda.gov/drugsatfda_docs/label/2023/208558s025lbl.pdf"

},

"Treatment 4": {

"Disease Name": "Prostate Cancer",

"Disease Phase or Condition": "Metastatic Castration-Resistant",

"Drug Name": "Abiraterone acetate + Niraparib",

"Prior Treatment or Resistance Status": "Not specified",

"Genomic Features": "Somatic variant in gene BRCA1",

"Link to FDA-approved Label": "https://www.accessdata.fda.gov/drugsatfda_docs/label/2023/216793s000lbl.pdf"

}

}

Supplementary Note 3. Example of a paired prompt-output from an actual physician’s query

**Prompt:**

Context information is below.

---------------------

['If a anaplastic large cell lymphoma (alcl) patient has a rearrangement in gene ALK, one recommended treatment would be Crizotinib. The U.S. Food and Drug Administration (FDA) granted approval to crizotinib for the treatment of pediatric patients 1 year of age or older and young adults with relapsed or refactory, systemic anaplastic large cell lymphoma (ALCL) that is ALK-positive. The safety and efficacy of crizotinib have not been established in older adults with relapsed or refactory systemic ALK-positive ALCL. (Citation: Pfizer, Inc. Xalkori (crizotinib) [package insert]. U.S. Food and Drug Administration website. https://www.accessdata.fda.gov/drugsatfda_docs/label/2022/202570s033lbl.pdf. Revised July 2022. Accessed August 4th, 2022.).', 'If a inflammatory myofibroblastic tumor (imt) patient has a rearrangement in gene ALK, one recommended treatment would be Crizotinib. The U.S. Food and Drug Administration (FDA) granted approval to crizotinib for the treatment of adult and pediatric patients 1 year of age or older with unresectable, recurrent, or refactory inflammatory anaplastic lymphoma kinase (ALK)-positive myofibroblastic tumors (IMT). (Citation: Pfizer, Inc. Xalkori (crizotinib) [package insert]. U.S. Food and Drug Administration website. https://www.accessdata.fda.gov/drugsatfda_docs/label/2022/202570s033lbl.pdf. Revised July 2022. Accessed August 4th, 2022.).', 'If a non-small cell lung cancer patient has a rearrangement in gene ALK, and is metastatic, one recommended treatment would be Ceritinib. The U.S. Food and Drug Administration (FDA) granted regular approval for ceritinib for patients with for patients with metastatic non-small cell lung cancer (NSCLC) whose tumors are anaplastic lymphoma kinase (ALK)-positive as detected by an FDA-approved test. (Citation: Novartis Pharmaceuticals Corporation. Zykadia (ceritinib) [package insert]. U.S. Food and Drug Administration website. https://www.accessdata.fda.gov/drugsatfda_docs/label/2019/211225s000lbl.pdf. Revised March 2019. Accessed November 12, 2020.).', 'If a non-small cell lung cancer patient has a rearrangement in gene ALK, and is metastatic, one recommended treatment would be Crizotinib. The U.S. Food and Drug Administration (FDA) granted approval for crizotinib for the treatment of patients with ALK positive metastatic non-small cell lung cancer (NSCLC) as detected by an FDA-approved test. (Citation: Pfizer, Inc. Xalkori (crizotinib) [package insert]. U.S. Food and Drug Administration website. https://www.accessdata.fda.gov/drugsatfda_docs/label/2022/202570s033lbl.pdf. Revised July 2022. Accessed August 4th, 2022.).', 'If a any solid tumor patient has a rearrangement in gene NTRK2, one recommended treatment would be Larotrectinib. The U.S. Food and Drug Administration (FDA) granted approval for larotrectinib for the treatment of adult and pediatric with solid tumors that have NTRK gene fusions without a known resistance mechanism, are metastatic or surgical resection is likely to result in severe morbidity, and have no satisfactory alternative treatments or have progressed following treatment. Continued approval for this indication may be contingent upon veritifcation and description of clinical benefit in a confirmatory trials. (Citation: Loxo Oncology, Inc. Vitrakvi (larotrectinib) [package insert]. U.S. Food and Drug Administration website. https://www.accessdata.fda.gov/drugsatfda_docs/label/2018/210861s000lbl.pdf. Revised November 2018. Accessed November 12, 2020.).', 'If a non-small cell lung cancer patient has a rearrangement in gene ALK, and is metastatic, one recommended treatment would be Lorlatinib. The U.S. Food and Drug Administration (FDA) granted approval for lorbrena for patients with ALK positive metastatic non-small cell lung cancer (NSCLC) for second- or third-line treatment. Continued approval for this indication may be contingent upon veritifcation and description of clinical benefit in a confirmatory trial. (Citation: Pfizer, Inc. Lorbrena (lorlatinib) [package insert]. U.S. Food and Drug Administration website. https://www.accessdata.fda.gov/drugsatfda_docs/label/2020/210868s001lbl.pdf. Revised May 2020. Accessed November 12, 2020.).', 'If a any solid tumor patient has a rearrangement in gene NTRK1, one recommended treatment would be Larotrectinib. The U.S. Food and Drug Administration (FDA) granted approval for larotrectinib for the treatment of adult and pediatric with solid tumors that have NTRK gene fusions without a known resistance mechanism, are metastatic or surgical resection is likely to result in severe morbidity, and have no satisfactory alternative treatments or have progressed following treatment. Continued approval for this indication may be contingent upon veritifcation and description of clinical benefit in a confirmatory trials. (Citation: Loxo Oncology, Inc. Vitrakvi (larotrectinib) [package insert]. U.S. Food and Drug Administration website. https://www.accessdata.fda.gov/drugsatfda_docs/label/2018/210861s000lbl.pdf. Revised November 2018. Accessed November 12, 2020.).', 'If a non-small cell lung cancer patient has a rearrangement in gene ALK, and is metastatic, one recommended treatment would be Alectinib. The U.S. Food and Drug Administration (FDA) granted regular approval to alectinib for treatment of patients with anaplastic lymphoma kinase (ALK)-positive metastatic non-small cell lung cancer (NSCLC), as detected by an FDA-approved test. (Citation: Genentech, Inc. Alecensa (alectinib) [package insert]. U.S. Food and Drug Administration website. https://www.accessdata.fda.gov/drugsatfda_docs/label/2018/208434s004lbl.pdf. Revised June 2018. Accessed November 12, 2020.).', 'If a any solid tumor patient has a rearrangement in gene NTRK2, one recommended treatment would be Entrectinib. The U.S. Food and Drug Administration (FDA) granted accelerated approval for entrectinib for adult and pediatric patients older than 1 month with solid tumors that have a NTRK gene fusion without a known acquired resistance mutation, are metastatic or where surgical resection is likely to result in severe morbidity, and have progressed following treatment or have no satisfactory standard therapy. (Citation: Genentech, Inc. Rozlytrek (entrectinib) [package insert]. U.S. Food and Drug Administration website. https://www.accessdata.fda.gov/drugsatfda_docs/label/2023/212725Orig1s009lbl.pdf. Revised October 2023. Accessed November 1, 2023.).', 'If a cholangiocarcinoma patient has a rearrangement in gene FGFR2, one recommended treatment would be Pemigatinib. The U.S. Food and Drug Administraton (FDA) granted accelerated approval to Pemazyre (pemigatinib). Pemigatinib is a kinase inhibitor indicated for the treatment of adults with previously treated, unresectable locally advanced or metastatic cholangiocarcinoma with a fibroblast growth factor 2 (FGFR2) fusion or rearrangement as detected by an FDA-approved test. (Citation: Incyte Corporation. Pemazyre (pemigatinib) [package insert]. U.S. Food and Drug Administration website. www.accessdata.fda.gov/drugsatfda_docs/label/2022/213736s002lbl.pdf. Revised August 2022. Accessed September 8th, 2022.).']

---------------------

If no FDA-approved drugs are found, respond: "There are no FDA-approved drugs for the provided context."

If FDA-approved drugs are found, please provide each treatment as a json format with the following JSON schema.

{

"Treatment 1": {

"Disease Name": ,

"Disease Phase or Condition": ,

"Drug Name": ,

"Prior Treatment or Resistance Status": ,

"Genomic Features":

"FDA-approval status":

"Link to FDA-approved Label":

}

}

Query: Are ALK inhibitors approved for use in TFCP2 fusion positive rhabdomyosarcoma?

**Output:**

{

"Treatment 1": {

"Disease Name": "Anaplastic Large Cell Lymphoma (ALCL)",

"Disease Phase or Condition": "Relapsed or refractory, systemic",

"Drug Name": "Crizotinib",

"Prior Treatment or Resistance Status": "Relapsed or refractory",

"Genomic Features": "ALK-positive",

"FDA-approval status": "Approved",

"Link to FDA-approved Label": "https://www.accessdata.fda.gov/drugsatfda_docs/label/2022/202570s033lbl.pdf"

},

"Treatment 2": {

"Disease Name": "Inflammatory Myofibroblastic Tumor (IMT)",

"Disease Phase or Condition": "Unresectable, recurrent, or refractory",

"Drug Name": "Crizotinib",

"Prior Treatment or Resistance Status": "Unresectable, recurrent, or refractory",

"Genomic Features": "ALK-positive",

"FDA-approval status": "Approved",

"Link to FDA-approved Label": "https://www.accessdata.fda.gov/drugsatfda_docs/label/2022/202570s033lbl.pdf"

},

"Treatment 3": {

"Disease Name": "Non-Small Cell Lung Cancer (NSCLC)",

"Disease Phase or Condition": "Metastatic",

"Drug Name": "Ceritinib",

"Prior Treatment or Resistance Status": "None specified",

"Genomic Features": "ALK-positive",

"FDA-approval status": "Approved",

"Link to FDA-approved Label": "https://www.accessdata.fda.gov/drugsatfda_docs/label/2019/211225s000lbl.pdf"

},

"Treatment 4": {

"Disease Name": "Non-Small Cell Lung Cancer (NSCLC)",

"Disease Phase or Condition": "Metastatic",

"Drug Name": "Crizotinib",

"Prior Treatment or Resistance Status": "None specified",

"Genomic Features": "ALK-positive",

"FDA-approval status": "Approved",

"Link to FDA-approved Label": "https://www.accessdata.fda.gov/drugsatfda_docs/label/2022/202570s033lbl.pdf"

},

"Treatment 5": {

"Disease Name": "Non-Small Cell Lung Cancer (NSCLC)",

"Disease Phase or Condition": "Metastatic",

"Drug Name": "Lorlatinib",

"Prior Treatment or Resistance Status": "Second- or third-line treatment",

"Genomic Features": "ALK-positive",

"FDA-approval status": "Approved",

"Link to FDA-approved Label": "https://www.accessdata.fda.gov/drugsatfda_docs/label/2020/210868s001lbl.pdf"

},

"Treatment 6": {

"Disease Name": "Non-Small Cell Lung Cancer (NSCLC)",

"Disease Phase or Condition": "Metastatic",

"Drug Name": "Alectinib",

"Prior Treatment or Resistance Status": "None specified",

"Genomic Features": "ALK-positive",

"FDA-approval status": "Approved",

"Link to FDA-approved Label": "https://www.accessdata.fda.gov/drugsatfda_docs/label/2018/208434s004lbl.pdf"

}

}

Supplementary Figure

Supplementary Figure 1. Distribution of unstructured and structured datasets

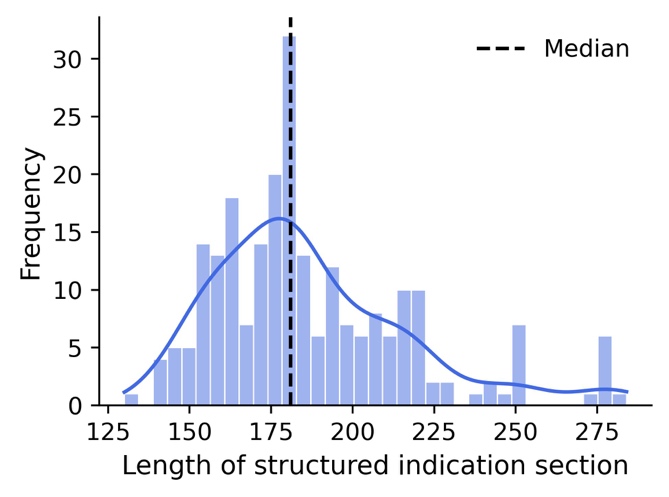

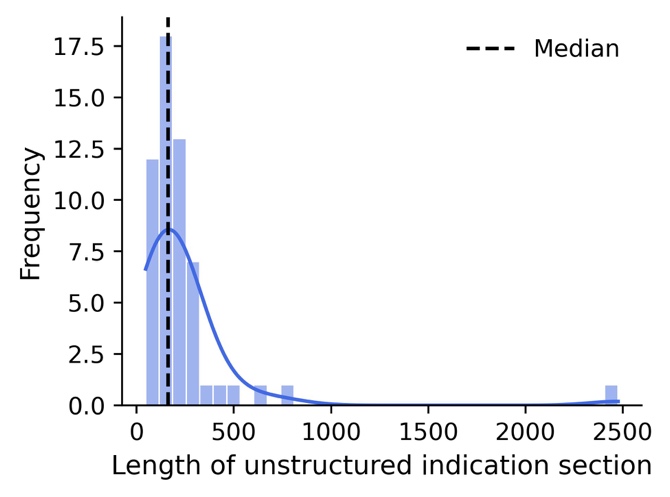

A

B

A. Distribution of the number of tokens in unstructured data directly extracted from the ‘Indication and Usage’ sections of FDA drug labels. B. Distribution of the number of tokens in structured data systematically curated, with each chunk representing a single use case derived from the structured MOAlamanc database.
